# Risk of acute gastroenteritis following COVID-19 exposure: a retrospective population-based analysis conducted in England

**DOI:** 10.64898/2026.09.07.26362439

**Authors:** Michael J. Hawkings, David M. Hughes, Alex J. Elliot, Iain Buchan, Daniel Hungerford

**Affiliations:** Department of Public Health Policy and Systems, University of Liverpool, L69 3GL, Liverpool, UK; NIHR Health Protection Research Unit in Gastrointestinal Infections at University of Liverpool, L697BE, Liverpool, UK; Department of Health Data Science, Institute of Population Health, University of Liverpool, Liverpool L69 3GF, UK; Civic Health Innovation Labs, University of Liverpool, Liverpool, UK; Real-time Syndromic Surveillance Team, Field Services, Health Protection in Regions, UK Health Security Agency, B2 4BH, Birmingham, UK; NIHR Health Protection Research Unit in Emerging and Zoonotic Infections at University of Liverpool, L697BE, Liverpool, UK; Department of Clinical Infection, Microbiology and Immunology, University of Liverpool, L697BE, Liverpool, UK

## Abstract

**Background:** Severe COVID-19 is associated with immune dysregulation, microbiome disruption, and increased vulnerability to secondary infections. However, the longer-term risk of acute gastroenteritis (AGE) following COVID-19 remains poorly studied. We compared AGE incidence among community-managed and hospitalised COVID-19 patients with test-negative controls and hospitalised sepsis patients.

**Methods:** We conducted a retrospective cohort study using electronic healthcare data. Patients with a recorded SARS-CoV-2 PCR test between 01 March 2020 and 01 March 2022 were allocated to test-negative, community COVID-19, or hospitalised COVID-19 cohorts. A hospitalised sepsis cohort was assembled for an age-, sex- and time-matched analysis. AGE incidence rates were calculated for the two years before enrolment and during follow-up. Negative binomial regression and conditional Poisson models estimated incidence rate ratios (IRRs) adjusting for demographic and clinical covariates.

**Results:** Among 788 103 patients, AGE incidence in the pre-pandemic period was higher among individuals later hospitalised with COVID-19 or sepsis compared with community cases. During follow-up, AGE incidence was slightly lower in community COVID-19 cases than test-negative controls (adjusted IRR 0·88 [95% CI 0·85–0·92]), but substantially higher among hospitalised COVID-19 patients (IRR 5·80 [95% CI: 5·38–6·25]). Matched analyses confirmed increased AGE rates compared with community controls (IRR 4·63 [95% CI: 4·15–5·17]) and hospitalised sepsis controls (IRR 1·41 [95% CI 1·21–1·64]).

**Conclusions:** COVID-19 hospitalisation is associated with a marked increase in AGE risk after discharge, exceeding that associated with sepsis hospitalisation. By contrast, lower AGE rates in community COVID-19 cases likely reflect baseline differences rather than protective effects of infection. Overall, these findings demonstrate heightened post-discharge vulnerability to gastrointestinal illness after COVID-19 hospitalisation and support the need for improved follow-up and surveillance strategies.

**Funding:** National Institute for Health Research

## Introduction

Since the emergence of SARS-CoV-2 in 2019, there is growing interest in the long-term health outcomes associated with COVID-19. Beyond post-COVID syndrome, a growing body of evidence suggests that COVID-19 infection may increase risks of gastrointestinal illnesses such as irritable bowel syndrome and inflammatory bowel disease^1^. Evidence from early studies also suggested a heightened susceptibility to secondary or opportunistic infections among patients with a history of COVID-19 illness^2^. One area that remains poorly studied is that of gastrointestinal (GI) infections, despite the known effects SARS-CoV-2 has on the GI system.

Several mechanisms explain how SARS-CoV-2 infection can negatively impact GI health. SARS-CoV-2 is known to have a negative effect on the gut microbiome – primarily a depletion of beneficial commensals which support functional immunity – both during acute COVID-19 illness and for many months after convalescence^3^. Disruption of the renin-angiotensin-aldosterone system occurs as a result of host binging to angiotensin-converting enzyme-2 (ACE-2) receptors, which are highly expressed throughout the GI tract^4^. The subsequent dysregulation of ACE-2 function has been shown to affect amino-acid metabolism and the microbiota, which in turn increases susceptibility to sequalae such as colitis^5^. Viral antigen persistence in gut epithelial cells has also been identified beyond recovery from clinical illness, and correlated with a persistent immune response^6,7^.

The literature examining risk of GI infection following COVID-19 is limited. A 2020 case report from Italy highlighted a case of reactivation of Giardiasis, an enteric parasitic disease with GI symptoms, in a 66 year-old male with a history of severe COVID-19. The authors suggested a mechanism of *Giardia* evading immune detection as a result of COVID-19 associated lymphopenia^8^. On the other hand, a 2023 Iranian study detected no evidence of rotavirus co-infection in stool samples collected from 37 children admitted to hospital with COVID-19^9^. Population-level data examined by Love et al. identified an overall net decrease in circulating gastrointestinal pathogens in the early stages of the COVID-19 pandemic^10^. This was attributed to public health and social measures (PHSMs), and changes to healthcare seeking behaviour, in addition to a true decrease in circulating pathogens, especially those where person-person transmission is the main mode of transmission. However, national surveillance data for England showed detections of common GI pathogens such as rotavirus and norovirus and syndromic consultations for gastroenteritis and diarrhoea had returned to pre-pandemic levels from summer 2021. There were also indications that syndromic AGE primary care attendances and norovirus detections were higher than pre-pandemic baseline levels in the years 2022 to 2024^11,12^.

Given the lack of high-quality epidemiological studies on the impact of COVID-19 infection on acute all-cause gastroenteritis (AGE), we conducted a population-based cohort study in England to estimate the differences in rates of acute gastroenteritis between COVID-19 negative, community and hospitalised patients.

## Methods

### Study design

We conducted a retrospective cohort study utilising routinely collected Electronic Health Record (EHR) data in a cohort of patients reporting a SARS-CoV-2 test result from 01 March 2020 to 01 March 2022 to investigate incident acute gastroenteritis. Data was available to 01 January 2015 to 31 December 2023. On reporting a SARS-CoV-2 test result, patients were allocated to one of three COVID-19 cohorts: test-negative, test-positive community, or COVID-19 hospitalised. We also assembled a control cohort of patients hospitalised with sepsis during the study period who had no COVID-19-related admissions.

### Source data

We utilised routine EHR data sourced from the Clinical Practice Research Datalink (CPRD). CPRD is an anonymised health record dataset compromising 18 million active patients, and considered the largest primary care database representative of the population of England in terms of age, sex and ethnicity^13^. For the purpose of our study, CPRD provides patient level primary care data individually linked to three additional healthcare datasets: (1) Second Generation Surveillance System (SGSS) laboratory data, (2) COVID-19 Hospitalisation in England Surveillance System (CHESS)/COVID-19 SARI-Watch hospitalisation data and, (3) Hospital Episode Statistics Admitted Patient Care (HES APC) data. We used the September 2024 build of CPRD Aurum for our study.

### Participants and variables

Contributing patients with confirmed linkage to the additional datasets were eligible for inclusion. COVID-19 status was ascertained through codes relating to PCR testing in either the primary care or linked SGSS dataset. Hospitalised patients were identified as those with linked HES APC or CHESS/COVID-19 SARI-Watch records who were admitted to hospital with a diagnosis of COVID-19 identified using the International Statistical Classification of Diseases and Related Health Problems 10th Revision codes (ICD-10) U07·1, U07·2 or U10·9 during the study period, including intensive care or high dependency units (ITU/HDU), and excluding patients who died during admission. COVID-19 status was treated as a time-varying exposure variable.

To ascertain AGE events, we utilised a code list established by Alexandridou et al. to capture ‘all-cause’ gastroenteritis^14^. This includes MEDCODE coded AGE events in primary care and ICD-10 coded events in secondary care. Our case definition of AGE was a single healthcare contact (limited to one per day) with no washout period; we selected this approach given the short duration of illness for the majority of GI infections. We formed our control cohort from ICD-10–coded admissions for sepsis and septic shock (A40, A41, R65·20, R65·21 and R57·2), capturing a disease severity profile comparable to that of COVID-19.

Vaccination against SARS-CoV-2 was ascertained by either a clinical event code or prescription issue within CPRD, with multiple events occurring within 60 days counted as one vaccination episode^15^. Patient comorbidity was measured using the validated 20-score Cambridge Multimorbidity Score (CMS)^16^. Socioeconomic status was defined by the patient-level English Index of Multiple Deprivation (IMD) 2019 quintile^17^. Patients were classified at baseline as “never smoked” and “history of smoking”. Code lists relating to covariates, including ethnicity, were constructed from various validated open sources and are presented in Appendix 1.

### Statistical analysis

Statistical analysis used R version 4·2·2. We initially produced a time-series of monthly AGE cases across primary and secondary care from January 2015 to December 2023. The crude incidence rate of AGE in the follow-up period and two years prior to enrolment was then calculated for COVID-19 negative, COVID-19 community, COVID-19 hospitalised and hospitalised sepsis patients.

We assessed overdispersion with the Poisson dispersion statistic. Based on this measurement, we fitted a negative binomial (NB) regression model, and tested whether a zero-inflated model added any additional value using the Vuong’s test. Incidence rate ratios (IRRs) were then estimated for the AGE during the follow up period. The initial multivariable model included age, sex, CMS score, IMD decile, smoking status, vaccination prior to exposure, and vaccination during the study period as explanatory covariates.

To account for the increased risk of AGE due to hospitalisation itself, we implemented a three-month lag on our follow-up period for patients with a valid discharge date in the HES APC or CHESS dataset. Secondly, we fitted a fixed-effects conditional Poisson model to estimate the IRR for AGE in a matched analysis; we paired one hospitalised COVID-19 patient with a community control, stipulating an exact match on age and CMS score, to account for age and comorbidity-associated confounding. Finally, we conducted an analysis of sepsis and COVID-19 hospitalised pairs using a fixed-effects conditional Poisson model. We applied coarsened exact matching using 10-year age bands, sex, and admission date (+/- 92 days).

## Results

From a cohort of 800,000 patients, we identified 788 103 participants with complete data relating to comorbidity, ethnicity, socioeconomic status and smoking status (Table 1), and following exclusion of patients who died during their hospital admission. Of these, 519 179 patients were allocated to our test-negative control cohort, 245 155 were allocated to our COVID-19 community cohort, and 23 769 allocated to our COVID-19 hospitalised cohort. 16 233 patients were allocated to our sepsis control cohort. Vaccine uptake during the study period was similar to levels reported nationally in the UK, however vaccination prior to exposure was much lower. Baseline comorbidity and mean age was highest amongst patients in both our COVID-19 and sepsis hospitalised cohorts.

**Table 1.**
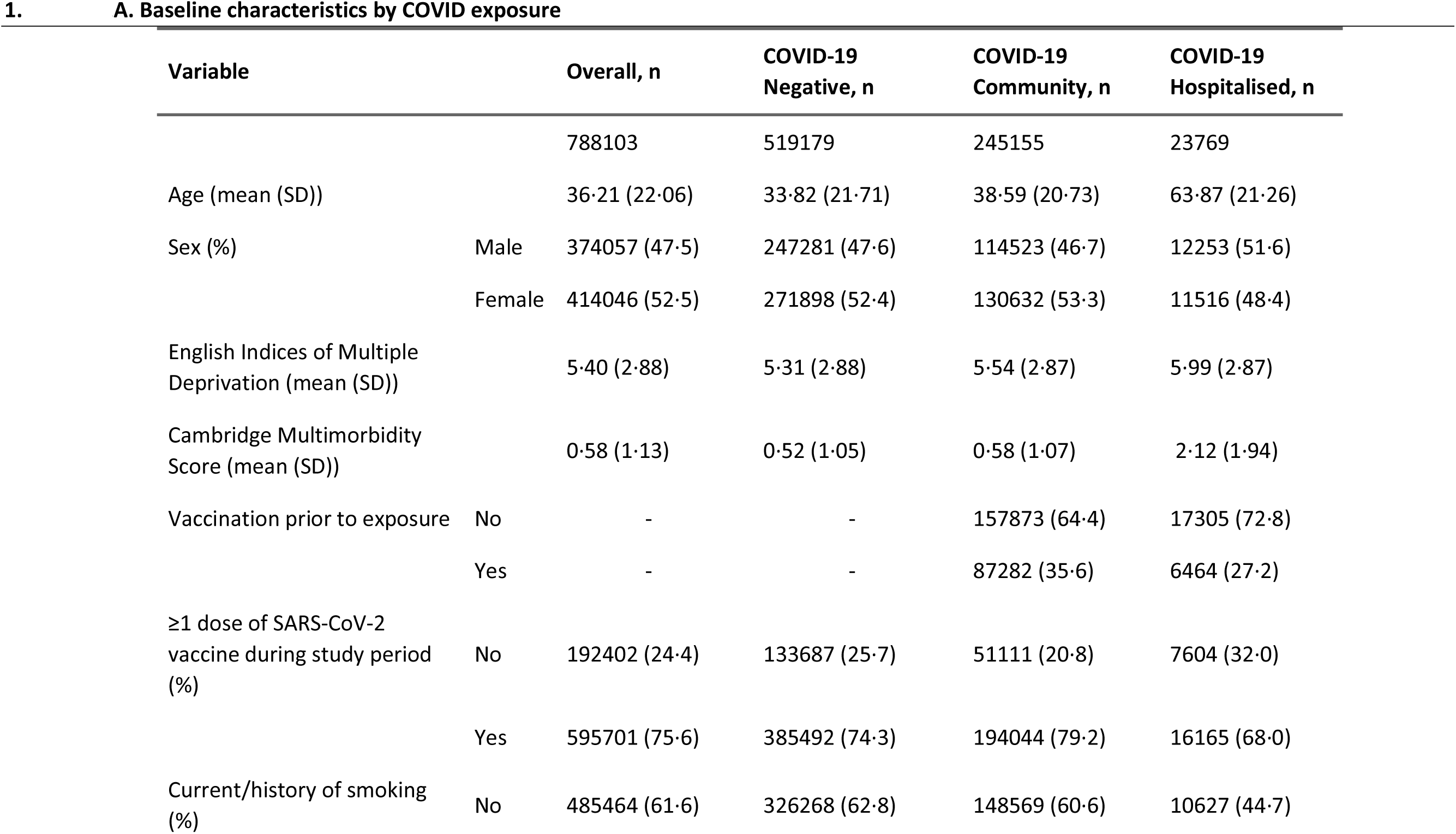
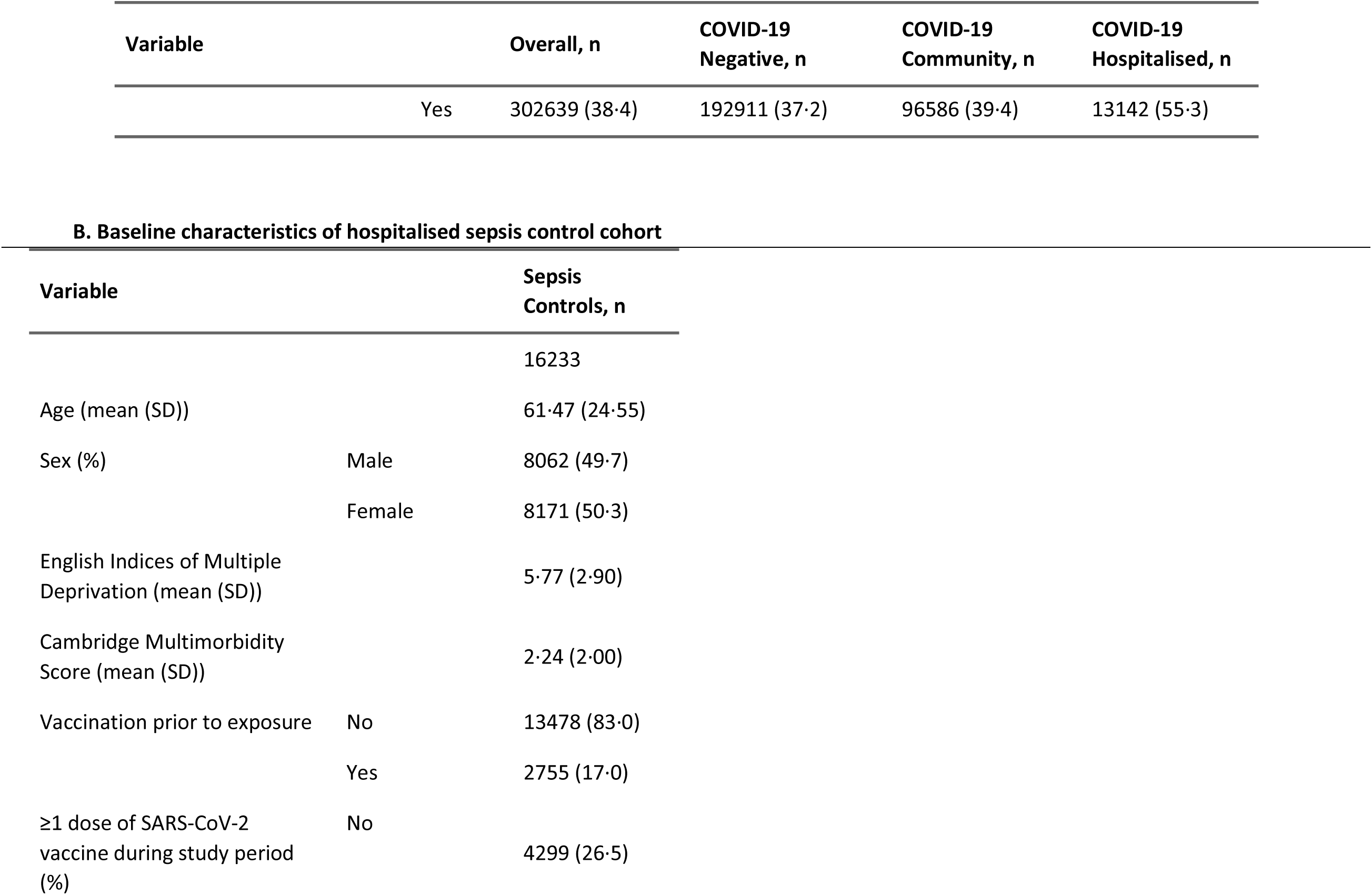
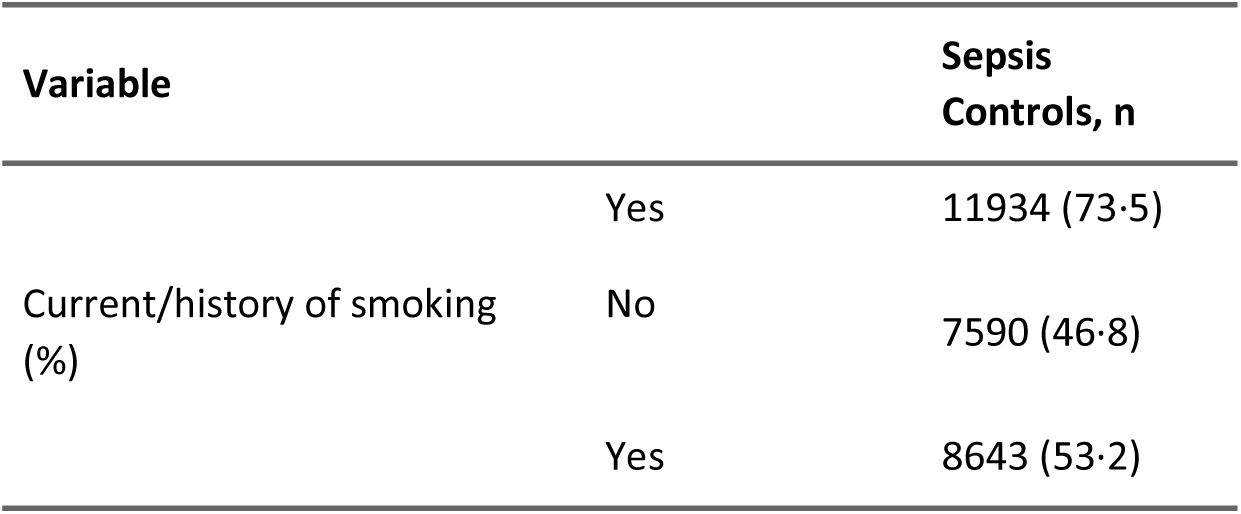
Baseline characteristics of entire cohort.

In the two years preceding enrolment, patients who were subsequently hospitalised with COVID-19 or sepsis had higher AGE incidence than our community cohort, with the highest rates observed among those later admitted for sepsis (Table 2). The incidence rate of AGE during the follow-up period was significantly higher for patients with a history of both COVID-19 and sepsis hospitalisation (Table 3). Furthermore, we observed a decrease in the incidence rate of AGE across primary and secondary care post-January 2020 during the early stages of the pandemic, however this began to returned to pre-pandemic levels towards the end of 2020 (Figure 1).

**Figure 1.**
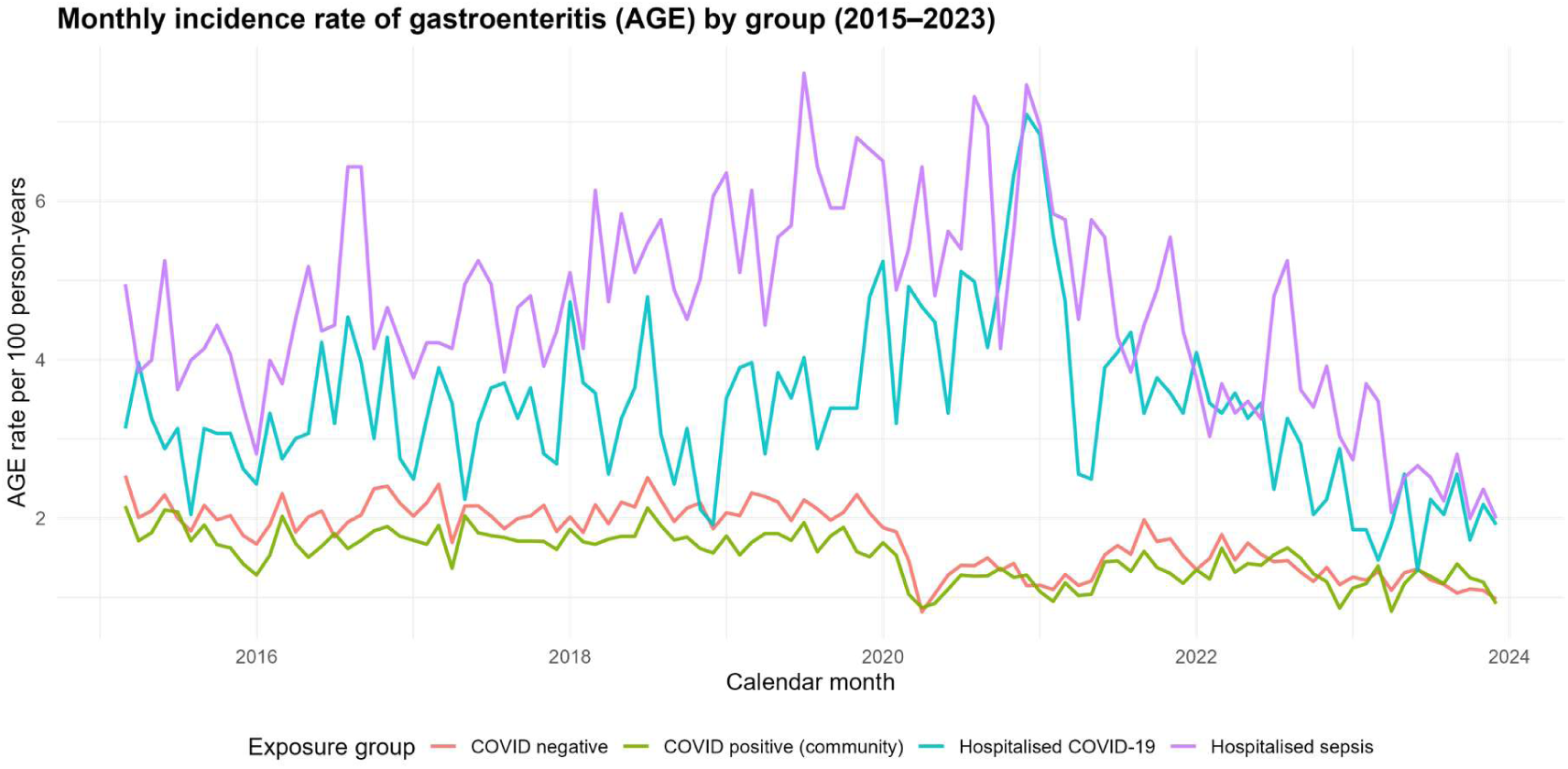
Time series showing monthly incidence of AGE in primary and secondary care by exposure group.

**Table 2.**
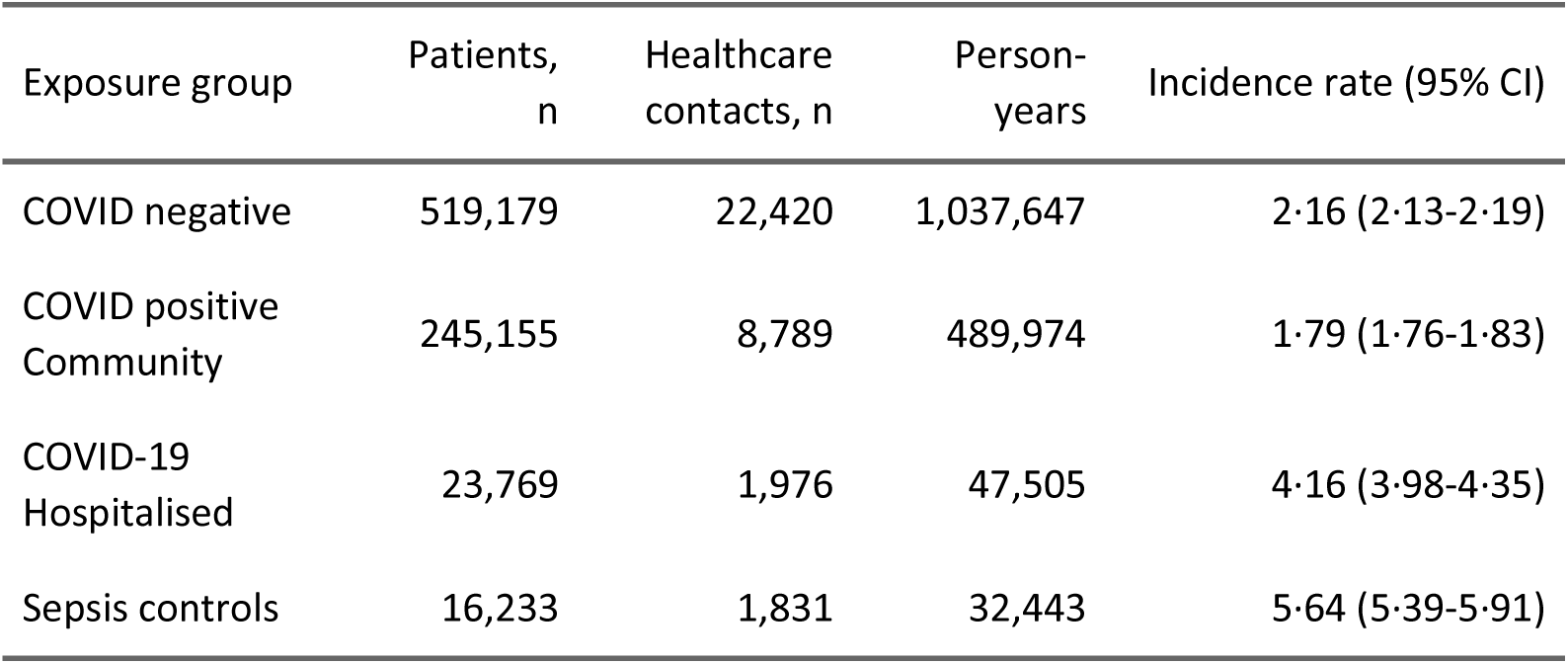
Incidence rate of AGE by exposure group per 100 person-years from 01 Jan 2018 to 31 Dec 2019.

**Table 3.** Incidence rate of AGE by exposure group per 100 person-years during the study period.

| Exposure group | Patients, n | Healthcare contacts, n | Person-years | Incidence rate (95% CI) |
| --- | --- | --- | --- | --- |
| COVID negative | 519,179 | 21,287 | 1,472,606 | 1·45 (1·43-1·47) |
| COVID positive Community | 245,155 | 9,092 | 695,360 | 1·31 (1·28-1·33) |
| COVID-19 Hospitalised | 23,769 | 2,979 | 67,418 | 4·42 (4·26-4·58) |
| Sepsis controls | 16,233 | 2,238 | 46,043 | 4·86 (4·66-5·07) |

For our univariable and multivariable models, the Poisson dispersion statistic was 2·45, favouring use of a negative binomial regression model. Vuong’s test showed no improved fit when using a zero-inflation component, so a negative binomial model was selected for our analysis. IRRs for our outcome and explanatory variables are presented in table 3.

**Table 3a.**
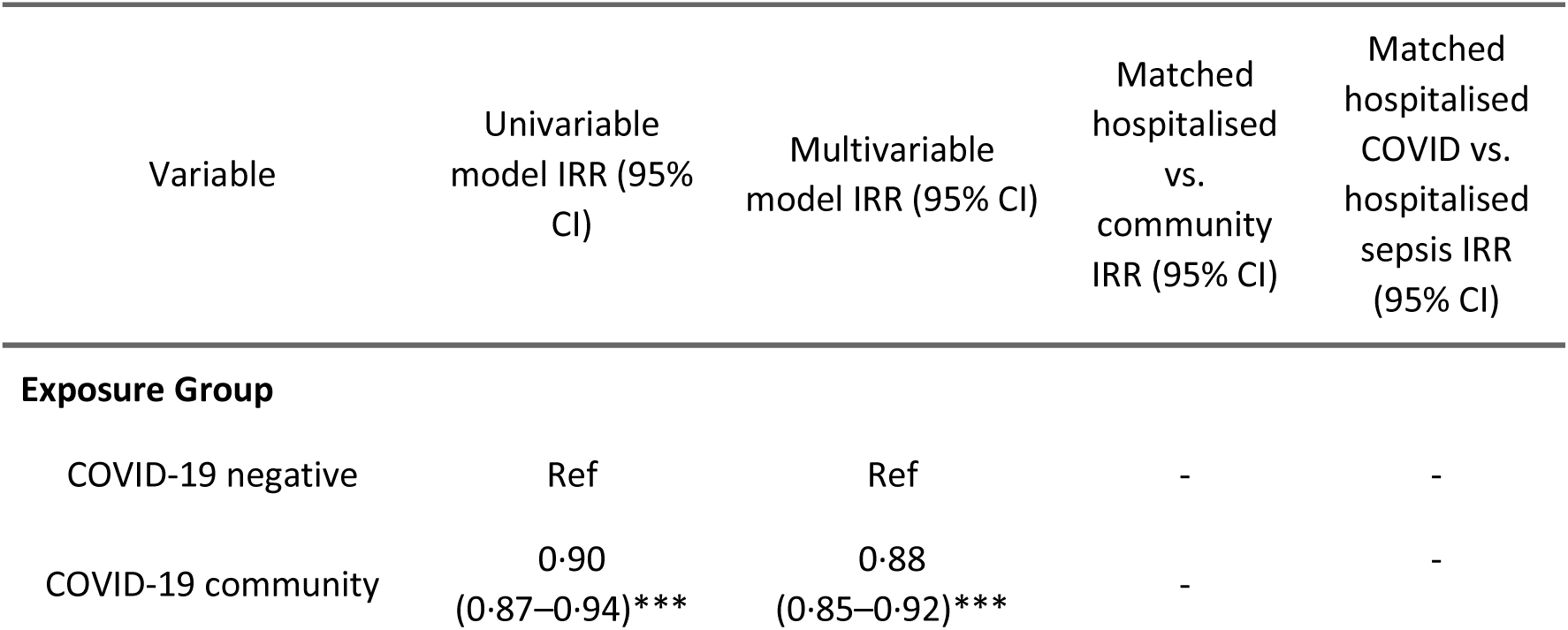
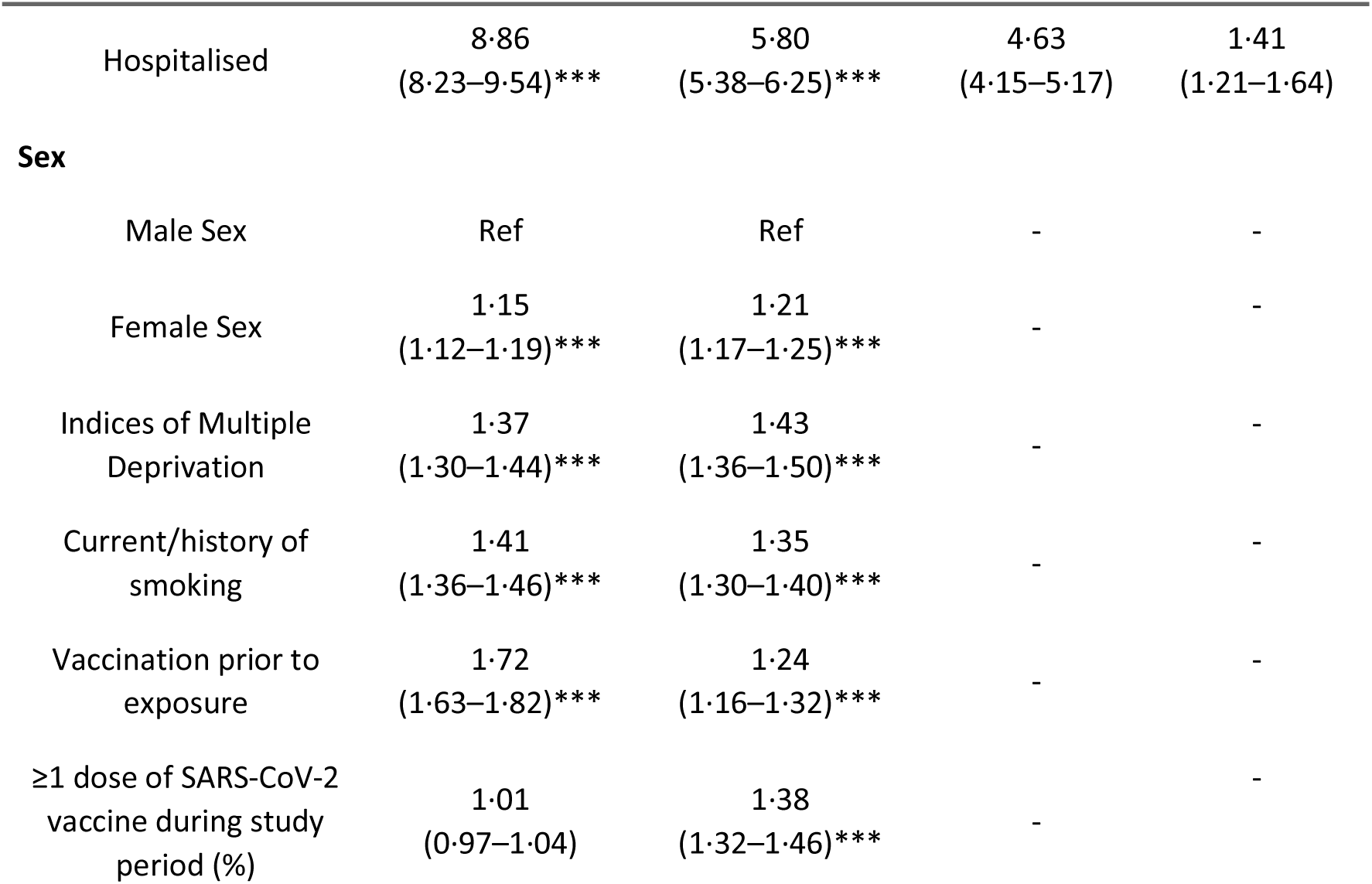
Incidence rate-ratios (IRRs) showing associations between study variables and AGE in primary and secondary care in a univariable model, multivariable model and a matched analysis for hospitalised vs community patients. *** P=<0·0001

In an analysis of 21 122 hospitalised and community pairs (42 244 patients), exactly matched on age and CMS score and followed for three months post discharge, hospitalisation with COVID-19 was associated with a markedly increased incidence of AGE (IRR 4·63, 95% CI 4·15–5·17) in a conditional fixed-effects Poisson model. In a separate analysis of 4 944 hospitalised COVID-19 and sepsis pairs (9 504 patients), exactly matched on 10-year age band, sex, and time of admission, COVID-19 hospitalisation was associated with higher AGE incidence compared with sepsis (IRR 1·41, 95% CI 1·21–1·64).

## Discussion

Our data suggest an increased incidence of AGE amongst patients previously hospitalised with COVID-19. By contrast, we identified a slightly decreased rate of AGE amongst COVID-19 cases managed in the community. This pattern was evident even in the years prior to the COVID-19 pandemic, indicating that the reduced AGE rate among community cases may be driven by pre-existing differences in baseline characteristics rather than by SARS-CoV-2 infection itself. Female sex, a current or past smoking history, and higher levels of socioeconomic deprivation were also associated with a modestly increased rate of AGE. We also observed a decrease in the overall incidence of AGE consultations across primary and secondary care, consistent with a decrease in healthcare utilisation, from early 2020 onwards. This returned to pre-pandemic levels by the beginning of 2021, consistent with national surveillance reports.

Hospitalisation, regardless of cause, is associated with an increased risk of secondary opportunistic infections, known as hospital-acquired/associated infection (HCAI). Specifically, patients with severe COVID-19, acute respiratory distress syndrome (ARDS) or sepsis are at an increased risk of HAI, including those affecting the GI tract^18^. Indeed, this is a leading cause of mortality amongst hospitalised COVID-19 patients^2^. It is also suggested, however, that surviving patients may enter a protracted immunosuppressive phase for many months, following the initial hyper-inflammatory phase associated with sepsis, severe infection and ARDS. This immune-dysregulation post-discharge may account for the increased susceptibility to AGE observed in our data, in addition to the known effects of SARS-CoV-2 on GI health^18^. We accounted for the possibility of hospital-acquired AGE presenting in our follow-up data by implementing a three-month diagnostic lag post-discharge, which resulted in consistent findings. Given gut microbiome dysbiosis and low-grade inflammation is known to persist for up to 12 months post-COVID-19, it may be that moderate-to-severe COVID-19 illness carries an increased risk for AGE independent of the immediate risk elevation associated with hospitalisation itself.

In the U.K., legislation introduced in early 2020 stipulated mandatory isolation for both individuals testing positive for SARS-CoV-2 and their household members. As a result, the modest protective effect for AGE observed amongst community COVID-19 cases is perhaps attributable to these patients being isolated from other pathogens associated with AGE. On the other hand, a proportion of patients previously hospitalised with COVID-19 may be discharged to care home settings, and be at an increased risk for AGE in the follow-up period as a result.

Vaccination prior to SARS-CoV-2 exposure was associated with increased recorded AGE incidence, likely reflecting residual confounding related to healthcare utilisation/healthy-user bias. In this study, vaccination was included as an adjustment covariate rather than modelled as an interaction term. We did not assess vaccination as an effect modifier of the association between COVID-19 exposure and AGE because AGE represents a broad, heterogeneous outcome with multiple infectious and non-infectious pathways, making interpretation of interaction effects challenging and potentially misleading. In addition, low numbers of individuals vaccinated prior to exposure, particularly among hospitalised patients, limited statistical power for stratified or interaction analyses.

### Strengths and limitations

This is the first study to examine the associations between a history of COVID-19 illness and the risk of subsequent AGE. This is in the context of well-established mechanisms highlighting the detrimental effects of SARS-CoV-2 on the GI system, and epidemiological studies suggesting GI sequalae to COVID-19. We utilised routine healthcare data that was representative of England, strengthened by robust case ascertainment offered by the availability of widespread SARS-CoV-2 testing early in the pandemic.

While biologically plausible, we are not able to attribute a direct causal relationship between SARS-CoV-2 and an increased risk of AGE from our observational data. This is due to both the differences in baseline characteristics between hospitalised and non-hospitalised COVID-19 patients, and the multitude of potential confounding factors associated with hospitalisation itself. To mitigate this, we assembled a control cohort of patients hospitalised with sepsis, matched on age, sex and admission date, and with no COVID-19–related admission. Sepsis was chosen given the similar clinical profile and spectrum of illness severity to that of COVID-19, and because UK data indicate high rates of antibiotic prescribing among COVID-19 inpatients^19,20^. Important differences remain, primarily the greater use of anticoagulants and immunomodulators in COVID-19 inpatients, which are not known to increase post-discharge AGE risk. Even within this hospitalised comparison, AGE incidence was higher in the COVID-19 cohort.

One of the strongest risk factors for enteric infection amongst hospitalised patients is proton pump inhibitor (PPI) exposure^21^. The use of broad-spectrum antibiotics in COVID-19 patients would also contribute to an increased risk for AGE^22^. While the majority of studies investigating AGE risk factors focus on the immediate susceptibility to GI infection amongst hospitalised patients, two studies suggest heightened risk for between one and three months post-cessation of PPI and antibiotic therapy^23,24^. While this was controlled for somewhat through matching to our antibiotic-exposed sepsis control cohort and the implementation of a three-month lag time, future studies should control for these time-varying confounders to better estimate the association between COVID-19, hospitalisation, and AGE.

Given we conducted a population-based study with community follow-up through GP records, it was not possible to estimate the incidence rate of *Clostridioides difficile* infections (CDI) in our cohort.

Previously, Granata et al. conducted a meta-analysis of incident infections in hospitalised COVID-19 patients and reported a pooled prevalence of 1% (95% CI: 1–2%) across seven studies^25^. Similarly, a large database cohort study of approximately one million hospitalised COVID-19 patients in the United States found that CDI co-infection was associated with longer hospital stays and higher mortality^26^. Given the likelihood that all-cause hospitalisation increases the risk of CDI, future studies should incorporate microbiology data to better identify specific pathogens contributing to the excess cases of AGE observed in our previously hospitalised cohort.

Our study suggests that patients previously exposed to SARS-CoV-2 may be at increased risk of GI morbidity, specifically an increased rate of primary and secondary care attendances for AGE post-COVID. Future studies would benefit from comparing AGE incidence between patients previously hospitalised with COVID-19 and a test-negative hospitalised control group, whilst accounting for different in-hospital treatments, to better estimate the effects of COVID-19 illness compared to hospitalisation itself.

## Data Availability

Data from this study is not available from the authors. Study data is available through formal request from CPRD. The datasets used in this study were extracted from CPRD following CPRD approval of the study protocol and through a data sharing agreement between University of Liverpool and CPRD. The same dataset is available from CPRD. The authors are not authorised to share the datasets and are obliged to destroy the datasets according to the data sharing agreement between University of Liverpool and CPRD

## Footnotes

## Acknowledgements

This study is based on data from the Clinical Practice Research Datalink (CPRD). CPRD is jointly sponsored by the Medicines and Healthcare products Regulatory Agency and the National Institute for Health Research (NIHR), as part of the Department of Health and Social Care. This work uses data provided by patients and collected by the National Health Service as part of their care and support. We would like to acknowledge all the data providers and general practices that made the anonymised data available for research. This study would not have been possible without the wider CPRD team at University of Liverpool including Pieta Schofield.

## Contributors

MJH, AJE, DH, IB, LB conceived of and designed the study. DH, MJH acquired the data; MJH analysed the data with support from DH, IB, DMH, and LB and all authors interpreted the output; MJH wrote the first draft of the report; and all authors reviewed the draft and final manuscript.

## Ethics approval and consent to participate

Ethical approval for this research was given by the Independent Scientific and Ethical Committee (ISAC) of the CPRD (reference 23_002987). CPRD obtains annual rolling ethical approval (reference number 05/MRE04/87) and no additional ethical approval was required for this project which meets ISAC requirements. The data was accessed via the University of Liverpool CPRD license agreement. All methods conformed to the ethical principles of the Declaration of Helsinki.

## Consent for publication

Not applicable

## References

1 Hawkings Michael J., Vaselli Natasha Marcella, Charalampopoulos Dimitrios, Brierley Liam, Elliot Alex J., Buchan Iain, et al. A Systematic Review of the Prevalence of Persistent Gastrointestinal Symptoms and Incidence of New Gastrointestinal Illness after Acute SARS-CoV-2 Infection. Viruses 2023;15(8):1625. Doi: 10.3390/v15081625.

2 Binkhamis Khalifa, Alhaider Alanoud S., Sayed Ayah K., Almufleh Yara K., Alarify Ghadah A., Alawlah Norah Y. Prevalence of secondary infections and association with mortality rates of hospitalized COVID-19 patients. Ann Saudi Med 2023;43(4):243–53. Doi: 10.5144/0256-4947.2023.243.

3 Righi Elda, Dalla Vecchia Ilaria, Auerbach Nina, Morra Matteo, Górska Anna, Sciammarella Concetta, et al. Gut Microbiome Disruption Following SARS-CoV-2: A Review. Microorganisms 2024;12(1):131. Doi: 10.3390/microorganisms12010131.

4 Qin W.-H., Liu C.-L., Jiang Y.-H., Hu B., Wang H.-Y., Fu J. Gut ACE2 Expression, Tryptophan Deficiency, and Inflammatory Responses The Potential Connection That Should Not Be Ignored During SARS-CoV-2 Infection. Cell Mol Gastroenterol Hepatol 2021;12(4):1514–1516.e4. Doi: 10.1016/j.jcmgh.2021.06.014.

5 Hashimoto Tatsuo, Perlot Thomas, Rehman Ateequr, Trichereau Jean, Ishiguro Hiroaki, Paolino Magdalena, et al. ACE2 links amino acid malnutrition to microbial ecology and intestinal inflammation. Nature 2012;487(7408):477–81. Doi: 10.1038/nature11228.

6 Zollner Andreas, Koch Robert, Jukic Almina, Pfister Alexandra, Meyer Moritz, Rössler Annika, et al. Postacute COVID-19 is Characterized by Gut Viral Antigen Persistence in Inflammatory Bowel Diseases. Gastroenterology 2022;163(2):495–506.e8. Doi: 10.1053/j.gastro.2022.04.037.

7 Zhou Yaya, Shi Xing, Fu Wei, Xiang Fei, He Xinliang, Yang Bohan, et al. Gut Microbiota Dysbiosis Correlates with Abnormal Immune Response in Moderate COVID-19 Patients with Fever. J Inflamm Res 2021;14:2619–31. Doi: 10.2147/JIR.S311518.

8 Lupia Tommaso, Corcione Silvia, De Rosa Francesco G. Giardiasis reactivation during severe SARS-CoV-2 infection. Parasitol Int 2021;80:102241. Doi: 10.1016/j.parint.2020.102241.

9 Zandi Milad, Soltani Saber, Sadooni Riam, Salmanzadeh Shokrollah, Erfani Yousef, Shahbahrami Ramin, et al. No sign of Rotavirus co-infection in COVID-19 patients with gastrointestinal symptoms. Malawi Med J 2023;35(1):27–30. Doi: 10.4314/mmj.v35i1.6.

10 Love Nicola K., Elliot Alex J., Chalmers Rachel M., Douglas Amy, Gharbia Saheer, McCormick Jacquelyn, et al. Impact of the COVID-19 pandemic on gastrointestinal infection trends in England, February-July 2020. BMJ Open 2022;12(3):e050469. Doi: 10.1136/bmjopen-2021-050469.

11 UKHSA norovirus-bulletin-2021-to-2022-week-28.pdf n.d.

12 GP in-hours: weekly bulletins for 2023. GOV.UK. Available at https://www.gov.uk/government/publications/gp-in-hours-weekly-bulletins-for-2023. Accessed September 30, 2025, 2024.

13 Sanchez-Santos Maria T, Axson Eleanor L, Dedman Daniel, Delmestri Antonella. Data Resource Profile Update: CPRD GOLD. Int J Epidemiol 2025;54(4):dyaf077. Doi: 10.1093/ije/dyaf077.

14 Alexandridou Maria, Cattaert Tom, Verstraeten Thomas. Estimation of Risk of Death Attributable to Acute Gastroenteritis Not Caused by Clostridioides difficile Infection Among Hospitalized Adults in England. Clin Epidemiol 2021;13:309–15. Doi: 10.2147/CLEP.S296516.

15 Andersen Kathleen M, McGrath Leah J, Reimbaeva Maya, Mendes Diana, Nguyen Jennifer L, Rai Kiran K, et al. Persons diagnosed with COVID-19 in England in the Clinical Practice Research Datalink (CPRD): a cohort description. BMJ Open 2024;14(1):e073866. Doi: 10.1136/bmjopen-2023-073866.

16 Payne Rupert A., Mendonca Silvia C., Elliott Marc N., Saunders Catherine L., Edwards Duncan A., Marshall Martin, et al. Development and validation of the Cambridge Multimorbidity Score. CMAJ 2020;192(5):E107–14. Doi: 10.1503/cmaj.190757.

17 UK Government. The English Indices of Deprivation 2019 n.d.

18 Hotchkiss Richard S., Monneret Guillaume, Payen Didier. Sepsis-induced immunosuppression: from cellular dysfunctions to immunotherapy. Nat Rev Immunol 2013;13(12):862–74. Doi: 10.1038/nri3552.

19 Russell Clark D., Fairfield Cameron J., Drake Thomas M., Turtle Lance, Seaton R. Andrew, Wootton Dan G., et al. Co-infections, secondary infections, and antimicrobial use in patients hospitalised with COVID-19 during the first pandemic wave from the ISARIC WHO CCP-UK study: a multicentre, prospective cohort study. The Lancet Microbe 2021;2(8):e354–65. Doi: 10.1016/S2666-5247(21)00090-2.

20 Surviving Sepsis Campaign: guidelines on the management of critically ill adults with Coronavirus Disease 2019 (COVID-19) | Intensive Care Medicine. Available at https://link.springer.com/article/10.1007/s00134-020-06022-5. Accessed November 12, 2025, n.d.

21 Shanika Lelwala Guruge Thushani, Reynolds Andrew, Pattison Sharon, Braund Rhiannon. Proton pump inhibitor use: systematic review of global trends and practices. Eur J Clin Pharmacol 2023;79(9):1159–72. Doi: 10.1007/s00228-023-03534-z.

22 Webb Brandon J., Subramanian Aruna, Lopansri Bert, Goodman Bruce, Jones Peter Bjorn, Ferraro Jeffrey, et al. Antibiotic Exposure and Risk for Hospital-Associated Clostridioides difficile Infection. Antimicrob Agents Chemother 2020;64(4):e02169–19. Doi: 10.1128/AAC.02169-19.

23 Hensgens Marjolein P. M., Goorhuis Abraham, Dekkers Olaf M., Kuijper Ed J. Time interval of increased risk for Clostridium difficile infection after exposure to antibiotics. J Antimicrob Chemother 2012;67(3):742–8. Doi: 10.1093/jac/dkr508.

24 McDonald Emily G., Milligan Jonathon, Frenette Charles, Lee Todd C. Continuous Proton Pump Inhibitor Therapy and the Associated Risk of Recurrent Clostridium difficile Infection. JAMA Intern Med 2015;175(5):784–91. Doi: 10.1001/jamainternmed.2015.42.

25 Granata Guido, Petrosillo Nicola, Al Moghazi Samir, Caraffa Emanuela, Puro Vincenzo, Tillotson Glenn, et al. The burden of Clostridioides difficile infection in COVID-19 patients: A systematic review and meta-analysis. Anaerobe 2022;74:102484. Doi: 10.1016/j.anaerobe.2021.102484.

26 Deda Xheni, Elfert Khaled, Gandhi Mustafa, Malik Alexander, Elromisy Esraa, Guevara Nehemias, et al. Clostridioides difficile Infection in COVID-19 Hospitalized Patients: A Nationwide Analysis. Gastroenterology Res 2023;16(4):234–9. Doi: 10.14740/gr1639.

